# Self-reported exposure to open air burn pits is associated with higher cancer prevalence in US Veterans

**DOI:** 10.64898/2026.05.01.26351950

**Authors:** Darren E. Gemoets, James J.S. Norton, Russell Hardesty, Maithao Le

## Abstract

Open air burn pits were used extensively during military operations in Iraq and Afghanistan, potentially exposing millions of US Veterans to toxic airborne hazards. Many of the airborne toxins released have been shown to induce lung inflammation and lung injury and are mutagenic. This is the first large-scale study of associations between self-reported burn pit exposures and the development of cancer. Using data from the Airborne Hazards and Open Burn Pit Registry, we found that Veterans reporting burn pit exposures are associated with a higher odds of developing cancer. However, investigations into the development of specific type of cancer and into a burn pit exposure dose-response effect were inconclusive.

---

During deployment to theaters of military operations in Iraq and Afghanistan, millions of US Veterans were potentially exposed to toxic airborne hazards released from open-air burn pits (BPs) [1]. BPs were used extensively during these operations to dispose of military, industrial, and medical waste. Toxins released by these sources include particulate matter, polycyclic aromatic hydrocarbons, volatile organic compounds, toxic organic halogenated dioxins and furanes (dioxins), and heavy metals. These airborne toxins have been shown to induce inflammation and lung injury and are mutagenic [2].

To date, no large-scale studies have investigated associations between the development of cancer and cumulative BP exposures in Veterans. Existing studies either utilized small sample sizes [3] or BP exposures were inferred [4]. Demonstrating the existence of associations between BP exposures and cancer is an important first step towards the establishment of causal links between BP exposures and cancer.

We performed the first large-scale investigation of associations between BP exposures and cancer using data from the Airborne Hazards and Open Burn Pit Registry (AHOBPR). The AHOBPR is a congressionally mandated registry that enables Veterans to voluntarily self-report their BP exposures and other health-related data (e.g., history of cancer). See [5] for additional details on the AHOBPR.

Our copy of the AHOBPR dataset (extracted March 2024) contains responses from 454, 223 Veterans, with our analysis performed on 309, 826 Veterans with complete cancer, BP exposure and demographic data (see Figure A.1 for exclusion diagram). Veterans were assigned to Cancer (*n* = 19, 567) or no-Cancer (*n* = 290, 259) groups based on self-reported cancer in the AHOBPR. Cumulative BP exposures experienced by each Veteran were measured (in days) using Department of Defense (DoD) deployment records and the AHOBPR. The Veterans were categorized in terms of BP exposed vs not exposed, with BP exposure further stratified by exposure quartiles for interpretability (see Table 1). Logistic regression was used to compute unadjusted and adjusted (for age at survey submission, race, and sex) odds ratios (ORs). See Appendix B for additional study method details.

**Table 1:** Demographic characteristics of the sample categorized by BP exposure level. Age at AHOBPR survey completion is reported as mean (SD); other variables reported as counts (percents). Percents are relative to demographic category. The last two rows present unadjusted and adjusted (for sex, age at AHOBPR completion, and race) ORs with no BP exposure as the reference. Cumulative BP exposure categories are defined by quartiles as follows: minimal (1 ≤ *days <* 43), low (43 ≤ *days <* 112); moderate (112 *< days <* 237), high (*days >* 237)

|  | No BP | Minimal | Low | Moderate | High |
| --- | --- | --- | --- | --- | --- |
| Age | 38.6 (9.6) | 40.7 (10.0) | 40.7 (9.8) | 40.7 (9.5) | 41.0 (8.5) |
| Male | 3209 (1.2) | 64564 (23.4) | 68849 (25.0) | 68949 (25.0) | 69977 (25.4) |
| Female | 541 (1.7) | 7404 (24.0) | 8051 (26.1) | 7990 (25.9) | 6889 (22.3) |
| Unknown | 37 (1.1) | 703 (20.7) | 930 (27.3) | 818 (24.0) | 915 (26.9) |
| White | 2840 (1.2) | 54735 (23.9) | 57547 (25.2) | 57276 (25.0) | 56264 (24.6) |
| Black | 518 (1.2) | 9466 (21.7) | 11288 (25.9) | 11095 (25.4) | 11277 (25.8) |
| Other | 247 (1.2) | 4457 (21.7) | 4676 (22.7) | 4977 (24.2) | 6216 (30.2) |
| Unknown | 182 (1.1) | 4013 (23.7) | 4319 (25.5) | 4409 (26.0) | 4024 (23.7) |
| Unadj OR | ref | 1.43 (1.23,1.68) | 1.43 (1.22,1.67) | 1.46 (1.25,1.71) | 1.48 (1.27,1.73) |
| Adj OR | ref | 1.24 (1.05,1.46) | 1.27 (1.09,1.50) | 1.33 (1.14,1.57) | 1.41 (1.20,1.66) |

The final cohort contained 309, 826 Veterans with a mean (SD) age of 40.8 (9.5) years; they were 88.9% male, 73.8% white and 14.1% Black. The majority of the Veterans served in Kuwait (33.4%), Iraq (31.3%) and Afghanistan (22.1%). In the 306,039 Veterans exposed to BPs, the median exposure time was 112 days (IQR: 43 to 237 days). Demographic data by BP exposure level are reported in Table 1.

Veterans who reported BP exposures also reported cancer diagnoses at a higher rate (6.3%) than non-exposed Veterans (4.5%), resulting in a significant increase in odds of cancer in Veterans exposed to BPs (OR 1.45, 95% CI: (1.25, 1.70)). The increased odds of cancer development persisted after adjustment for age, sex, and race (OR 1.31, 95% CI: (1.12,1.54)). However, our analyses did not find a BP exposure dose effect, evidenced by comparable odds ratios across exposure groups and overlapping confidence intervals; see Table 1.

Analyses of the data for associations between BP exposures and cancer types were inconclusive (analysis not shown). However, the distribution of cancer prevalence by type and BP exposure category are provided in Table C.2 in Appendix C.

For AHOBPR Veterans, we found that self-reported BP exposures are associated with a significantly higher odds of cancer development compared to Veterans without BP exposures. We did not find an exposure dose-effect. This may be due to the limited time of BP exposures (i.e., on the scale of days versus years, as for people with toxic exposures due to smoking). More analyses are needed to further evaluate potential BP exposure dose-effects and the effect of BP exposure on specific cancer types. Limitations of our study include self reported data inherent to a registry, and a proportionally small sample of non-BP-exposed Veterans. Strength of the study is the large sample size and granular data on Veterans BP exposures contained in the AHOBPR. The association between BP exposures and cancer development established in this study is an important first step towards the establishment of causal links between BP exposures and cancer and the development of targeted screening protocols for the early identification of BP-related cancers.

## Data Availability

The data used for this study are the property of the US Department of Veterans Affairs. The authors are unable to provide access to the data.

## Abbreviations

AHOBPR: Airborne Hazards and Open Burn Pit Registry
BP: burn pit
CI: confidence interval
DoD: Department of Defense
IQR: interquartile range
OR: odds ratio
SD: standard deviation

## Availability of data and materials

The data used for this study are the property of the US Department of Veterans Affairs. The authors are unable to provide data access.

## Acknowledgments

This material is the result of work supported with resources and the use of facilities at the Stratton VAMC, Albany, NY. Darren E. Gemoets, Ph.D. and Maithao Le, MD Ph.D. are employees of the US Department of Veterans Affairs and their opinions expressed in this paper are those of the authors’ and do not represent the views of the Department of Veterans Affairs or the US Government.

This work was supported using resources and facilities at the VA Informatics and Computing Infrastructure, VA HSR RES 13-457.

## Author Contributions

Darren E. Gemoets performed the data analysis and prepared early drafts of the manuscript. James J.S. Norton contributed to manuscript writing and data analysis. Russell Hardesty performed data cleaning and processing. Maithao Le conceptualized the project and provided clinical expertise. All authors contributed to finalizing the manuscript, and all approve the accuracy and integrity of the study.

## Appendix A Inclusion/exclusion diagram

**Figure A1:**
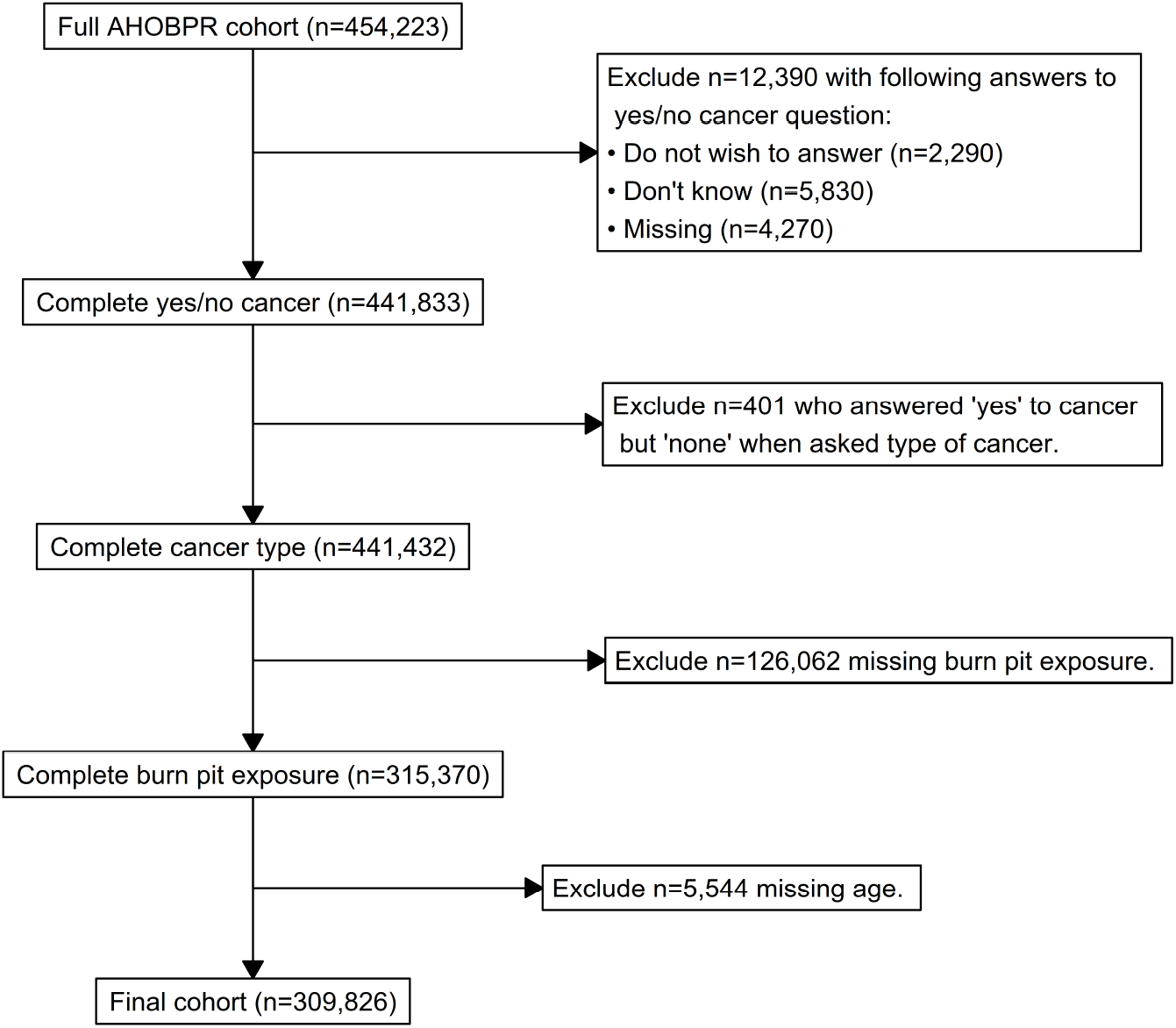
Diagram outlining the Veterans excluded from the study. The first stage excluded *n* = 12, 390 Veterans who did not provide a yes or no answer to the question “Have you **ever** been told by a doctor or other health professional that you had Cancer or malignancy (tumor) of any kind?”, with distribution of missing provided in the figure. The next stage excluded *n* = 410 Veterans who answered “yes” to the previous cancer question but “none” to the follow up question regarding type of cancer. The last two stages excluded *n* = 126, 062 Veterans who did not answer the question “On a typical day, how many hours did smoke or fumes from the burn pit enter your worksite or housing?” for their deployments, and *n* = 5, 544 who did not provide their current age.

## Appendix B Methods Miscellanea

### Study Design

Although we can infer and estimate BP exposure dates from DoD deployment dates from the AHOBPR, cancer diagnosis dates are not recorded. Thus we used of a cross sectional study design.

### Cancer variable

The phrasing of the question in the AHOBPR used to assigned Veterans to Cancer and no-Cancer groups is as follows, “Have you **ever** been told by a doctor or other health professional that you had Cancer or malignancy (tumor) of any kind?”. The possible answers are “Yes”, “No”, “Do not wish to answer”, “Don’t know” or the Veteran can leave it blank. Only Veterans with answers of “Yes” or “No” were included in the study.

### Calculation of burn pit exposure variable

For each DoD confirmed deployment, the AHOBPR asked Veterans to report: “On a typical day, how many hours did smoke or fumes from the burn pit enter your **worksite or housing**?” BP exposures for each deployment were calculated as the product of the length of the deployment (in days) and the typical number of hours of BP exposure that the Veteran experienced each day. The cumulative BP exposures experienced by each Veteran were then calculated by summing across deployments. We further categorized BP exposure using quartiles as follows: minimal (1 ≤ *days <* 43), low (43 ≤ *days <* 112); moderate (112 *< days <* 237), high (*days >* 237).

### Computational resources and software

All data analysis was performed using R (version 4.5.2) on the Veterans Affairs Informatics and Computing Infrastructure, which is a secure platform for research studies using Veterans Affairs electronic health record data.

### Approvals

Our study was approved by the Albany Stratton Veterans Affairs Medical Center Institutional Review Board (# 1334689).

## Appendix C Cancer prevalence by cancer type

**Table C.2:** Counts (%) of cancer cases by exposure category. Percents are calculated within rows.

| Cancer Type | No BPE | Minimal | Low | Moderate | High |
| --- | --- | --- | --- | --- | --- |
| Bladder | 0 (0) | 73 (24.7) | 85 (28.8) | 75 (25.4) | 62 (21) |
| Blood | 1 (0.5) | 53 (26.2) | 50 (24.8) | 50 (24.8) | 48 (23.8) |
| Bone | 0 (0) | 26 (19) | 39 (28.5) | 30 (21.9) | 42 (30.7) |
| Brain | 1 (0.3) | 70 (18.7) | 81 (21.7) | 121 (32.4) | 101 (27) |
| Breast | 7 (1.2) | 141 (24.3) | 145 (25) | 139 (23.9) | 149 (25.6) |
| Cervix | 2 (0.4) | 99 (19.6) | 126 (24.9) | 135 (26.7) | 144 (28.5) |
| Colon | 8 (1) | 175 (21.9) | 187 (23.4) | 211 (26.4) | 218 (27.3) |
| Esophagus | 1 (1.1) | 21 (22.1) | 25 (26.3) | 28 (29.5) | 20 (21.1) |
| Gallbladder | 0 (0) | 8 (36.4) | 5 (22.7) | 6 (27.3) | 3 (13.6) |
| Kidney | 4 (0.7) | 122 (22.8) | 113 (21.2) | 154 (28.8) | 141 (26.4) |
| Larynx-windpipe | 0 (0) | 13 (40.6) | 4 (12.5) | 6 (18.8) | 9 (28.1) |
| Leukemia | 4 (1.2) | 76 (22.8) | 81 (24.3) | 97 (29) | 76 (22.8) |
| Liver | 0 (0) | 20 (24.4) | 20 (24.4) | 20 (24.4) | 22 (26.8) |
| Lung | 0 (0) | 69 (22.3) | 91 (29.4) | 62 (20) | 88 (28.4) |
| Lymphoma | 9 (1.1) | 185 (22.6) | 215 (26.3) | 210 (25.7) | 199 (24.3) |
| Melanoma | 19 (0.9) | 494 (23.5) | 525 (25) | 552 (26.3) | 510 (24.3) |
| Mouth/tongue/lip | 3 (1.6) | 40 (22) | 43 (23.6) | 51 (28) | 45 (24.7) |
| Ovary | 1 (0.9) | 27 (24.1) | 33 (29.5) | 22 (19.6) | 29 (25.9) |
| Pancreas | 0 (0) | 17 (22.7) | 19 (25.3) | 17 (22.7) | 22 (29.3) |
| Prostate | 16 (1) | 359 (23.2) | 417 (26.9) | 410 (26.5) | 347 (22.4) |
| Rectum | 0 (0) | 28 (21.2) | 33 (25) | 43 (32.6) | 28 (21.2) |
| Skin (Don't know what kind) | 12 (0.6) | 459 (22.1) | 522 (25.1) | 501 (24.1) | 583 (28.1) |
| Skin (non-melanoma) | 41 (1) | 987 (25.2) | 964 (24.6) | 947 (24.2) | 974 (24.9) |
| Soft tissue (muscle or fat) | 4 (0.9) | 92 (20.4) | 96 (21.3) | 123 (27.3) | 136 (30.2) |
| Stomach | 1 (1.2) | 23 (27.4) | 16 (19) | 19 (22.6) | 25 (29.8) |
| Testis | 7 (0.8) | 172 (20.5) | 218 (26) | 216 (25.8) | 225 (26.8) |
| Throat-pharynx | 2 (1.1) | 41 (22.8) | 49 (27.2) | 41 (22.8) | 47 (26.1) |
| Thyroid | 5 (0.7) | 180 (23.9) | 168 (22.3) | 206 (27.4) | 193 (25.7) |
| Uterus | 1 (0.8) | 21 (16.9) | 29 (23.4) | 28 (22.6) | 45 (36.3) |

